# Cost-effectiveness analysis of a maternal pneumococcal vaccine in low-income, high-burden settings such as Sierra Leone

**DOI:** 10.1101/2022.07.25.22278016

**Authors:** Gizem M. Bilgin, Syarifah Liza Munira, Kamalini Lokuge, Kathryn Glass

## Abstract

Maternal pneumococcal vaccines have been proposed as a method of protecting infants in the first few months of life. In this paper, we assess the cost-effectiveness of a maternal pneumococcal polysaccharide vaccine using a health sector perspective. We estimate the costs of delivering a maternal pneumococcal polysaccharide vaccine, and the healthcare costs averted through the prevention of severe pneumococcal outcomes such as pneumonia and meningitis, with estimates of vaccine effectiveness based on previous research. Our model estimates that a maternal pneumococcal program would cost $570 per DALY averted (2020 USD, range $558-582), and hence not be cost-effective in our study setting of Sierra Leone using the nation’s GDP per capita of $527 as a benchmark. However, the choice of discounting rates for health outcomes determines whether the maternal pneumococcal vaccine was deemed cost-effective. Without discounting, the cost per DALY averted would be $277 (53% of Sierra Leone’s GDP per capita). Further, the cost per DALY averted would be $128 (24% GDP per capita) if PPV could be procured at the same cost relative to PCV in Sierra Leone as on the PAHO reference price list. Overall, our paper demonstrates that maternal pneumococcal vaccines have the potential to be cost-effective in low-income settings; however, their cost-effectiveness depends on the choice of discounting rates determined by social values, and negotiations with vaccine providers on vaccine price. Vaccine price is the largest cost driving the cost-effectiveness of a future maternal pneumococcal vaccine.

## 1. Introduction

The various presentations of pneumococcal disease, including pneumonia and meningitis, are a significant cause of morbidity and mortality in children worldwide [1]. The widespread adoption of pneumococcal conjugate vaccines (PCVs) has significantly reduced the burden of pneumococcal disease [2, 3]; however, the largest burden of pneumococcal disease is now in the first few months of life before childhood immunisations commence [4, 5].

Maternal vaccination with pneumococcal polysaccharide vaccines (PPVs) have been proposed as a strategy for protection in early childhood before PCV-derived immunity develops [6]. Maternal vaccines for influenza, tetanus and pertussis have already been demonstrated to provide protection to infants within their first few months of life [7-9]. To date, only preliminary randomised control trials and case-control studies of maternal PPV have been conducted [6]. These studies have observed no adverse outcomes of PPVs in pregnancy but have yielded inconclusive results regarding the efficacy of PPV as a maternal vaccine due to their small-scale. In a previous paper, we demonstrated that a maternal pneumococcal vaccine could reduce incidence by 73% (range 49-88%) in children <1 month, and 55% (range 36-66%) in children 1-2 months old [10].

This paper explores whether a maternal PPV could be cost-effective in reducing the burden of infant pneumococcal disease. We focus on modelling a low-income setting with a well-established infant PCV schedule, since maternal pneumococcal vaccination is primarily proposed as a supplement to protect the youngest infants in these settings.

## 2. Methods

### 2.1 Overview

In this study, we quantify the incremental cost-effectiveness of introducing a maternal PPV vaccine into a setting with existing infant PCV. The impacts of a maternal pneumococcal vaccine on health outcomes have been quantified in a previous paper [10]. Here, we estimate the costs of introducing a maternal pneumococcal vaccine, and the healthcare costs averted through the novel use of this vaccine. Our analyses adopt a health sector perspective and are presented for one hypothetical year of maternal vaccine delivery. Discounting was applied only to the derived metric of disability-adjusted life years (DALYs). All other health benefits and health costs occurred during the first year of life, so were not discounted. All prices were adjusted to 2020 United States Dollars (USD) using the International Monetary Fund’s gross domestic product (GDP) deflators [11] following methods proposed by Turner and colleagues [12].

### 2.2 Study setting

Sierra Leone was the primary setting chosen for the model, given it is a low-income country with a very high burden of pneumococcal disease in early childhood. It is a West African nation in Sub-Saharan Africa, the region where half of all pneumococcal-associated deaths in children under five occur today [1]. Although PCV has been a routine vaccine in Sierra Leone since 2011 [13], there remains a high rate of infant mortality with 75 deaths per 1,000 live births [14]. The government of Sierra Leone are committed to reducing deaths in children under five, having introduced a Free Health Care Initiative in 2010 that waives all medical fees for pregnant and breastfeeding women, and for children under 5 years of age [15]. There is high uptake of a maternal tetanus vaccine in Sierra Leone, with 97.4% of pregnant women receiving at least one dose during pregnancy [14]. A single dose of PPV is sufficient [16].

### 2.3 Health outcomes

#### 2.3.1 Disease model

The details of the disease model have been described in a previous paper [10]. In brief, we constructed a dynamic Susceptible-Infected-Suspectable (SIS) model. The model contained detailed age-structure for infants under two, with additional classes representing PCV-derived immunity. The model was fitted to the prevalence of pneumococcal-attributable acute respiratory illness. We introduced a maternal vaccine to this fitted model and used reductions in pneumococcal incidence to estimate reductions in pneumococcal-associated health outcomes. We considered the effects of introducing a maternal vaccine for children under one year of age since maternally derived immunity is expected to wane after one year [17]. No adverse events related to the vaccine were included in the model since such events have not been documented in existing trial data [6].

#### 2.3.2 Translation of health outcomes to DALYs

We translated estimates of health outcomes averted into disability-adjusted life years (DALYs) to align with previous cost-effectiveness analysis of pneumococcal vaccines [18-22]. We determined years of life lost (YLL) using life expectancy at birth from United Nations population estimates for 2020-2024 [23]. We calculated years lived with disability (YLD) using disability weights from the Global Burden of Disease Study 2016 [24]. We employed the ‘severe lower respiratory infection’ weight (0.133; 95% CI 0.088-0.190) for all non-fatal IPD episodes, and ‘moderate lower respiratory infection’ weight (0.051; 95% CI 0.032-0.074) for all non-invasive pneumococcal disease episodes. The duration of symptomatic respiratory infection was assumed to be 10 days (9-12 days) [24]. We assumed that 24.7% of all pneumococcal meningitis cases developed lifelong meningitis sequalae [25], and applied the disability weight for ‘severe motor plus cognitive impartments’ (0.542; 95% 0.374-0.702), as per Ojal et al. [21], to these episodes. A discounting rate of 3% was applied in line with existing cost-effectiveness analysis in Sub-Saharan Africa [19-22]. Discounting was applied using a continuous time approach with uniform age weighting [26].

### 2.4 Vaccine program costs

We split vaccine program costs into two components: vaccine costs, and operational costs (Table 1). Vaccine costs included the costs to procure the vaccine and related injection equipment. Operational costs encompassed supply chain costs and service delivery costs.

**Table 1.** Vaccine program costing parameters with point estimates, distributions, and sources. All prices are in 2020 USD.

| Parameter | Component description | Point estimate | Distribution | Reference |
| --- | --- | --- | --- | --- |
| Cost of vaccine | Price per dose | \$8.63 | Fixed | [27, 28] |
|  | Wastage rate | 5% | Fixed | [13, 29] |
|  | Freight costs | 4.5% of vaccine value | Fixed | [29] |
| Injection equipment | Bundled price for auto-disable (AD) syringes and safety boxes (per dose) | \$0.044 | Fixed | [30] |
|  | Wastage rate | 10% | Fixed | [29] |
| Operational costs | All non-vaccine and injections supply cost including the cost of personnel, training, transport, and social mobilisation (per dose) | \$0.97 | Gamma(6.25,0.15) | [31] |

#### 2.4.1 Vaccine costs

Vaccine costs included the price, wastage and freight costs of the vaccine, syringes, and safety boxes. PPVs have not been widely utilised in low-income nations since the introduction of PCV. The WHO’s 2020 Global Market Study for pneumococcal vaccines identified no low-income nation self-procuring PPV [32]. The study recognised that PPVs were primarily prescribed to adults over 65 and younger adults with comorbidities in high- and middle-income nations. Given the lack of a comparable setting, we estimated the price per dose of PPV from the Pan American Health Organisation’s (PAHO) vaccine price list for 2020 [28]. PAHO’s price per dose aligned with South Africa’s public health system price per dose of PPV (adjusted to 2020) [27]. Estimates of vaccine wastage and freight costs were taken from existing data on PCV. PPV wastage is expected to be low due to the vaccine’s presentation as a liquid single-dose vial.

Injection equipment costs were taken from United Nation Children’s Fund (UNICEF) Supply Division prices, as recommended in Sierra Leones’s Expanded Programme of Immunization Comprehensive Multi Year Plan (cMYP) [13, 30]. These costs aligned with previous in-country GAVI approved funding estimates for injection equipment [33]. UNICEF prices are listed as free carrier prices, meaning that they include freight costs.

#### 2.4.2 Operational costs

Operational costs included all non-vaccine and injection supply costs. That is, costs of transportation and storage, and costs of personnel to deliver the vaccine, program management and training. Operational costs for our hypothetical maternal pneumococcal vaccine were based on existing maternal tetanus vaccine data. The tetanus toxoid (TT) vaccine was the first maternal vaccine endorsed by the WHO and is the most widely used in low-income settings [34, 35]. Further, TT has similar temperature storage requirements to PPV [16, 36]. Sierra Leone’s latest cMYP does not include maternal tetanus vaccine specific operational costs [13]. Instead, our estimate of operational costs was informed by a global review of cMYPs submitted to WHO and UNICEF [31]. Notably, this global review’s estimates of operational costs for polio and measles aligned with the true operational costs for polio and measles reported in Sierra Leone [13, 31]. Further, the review’s estimate for maternal vaccine operational costs aligned with the operational costs per dose of maternal tetanus vaccination in Liberia, a country of similar size and neighbour to Sierra Leone [37]. We modelled the distribution of operational costs using a gamma distribution, as per previous cost-effectiveness analysis [21].

### 2.5 Health system costs averted

Pneumococcal cases were divided between invasive pneumococcal disease (IPD) and all other pneumococcal-attributable acute respiratory illnesses (ARI) (Fig 1). IPD presentations included pneumococcal pneumonia, pneumococcal meningitis, and non-pneumonia non-meningitis (NPNM) IPD.

**Fig 1.**
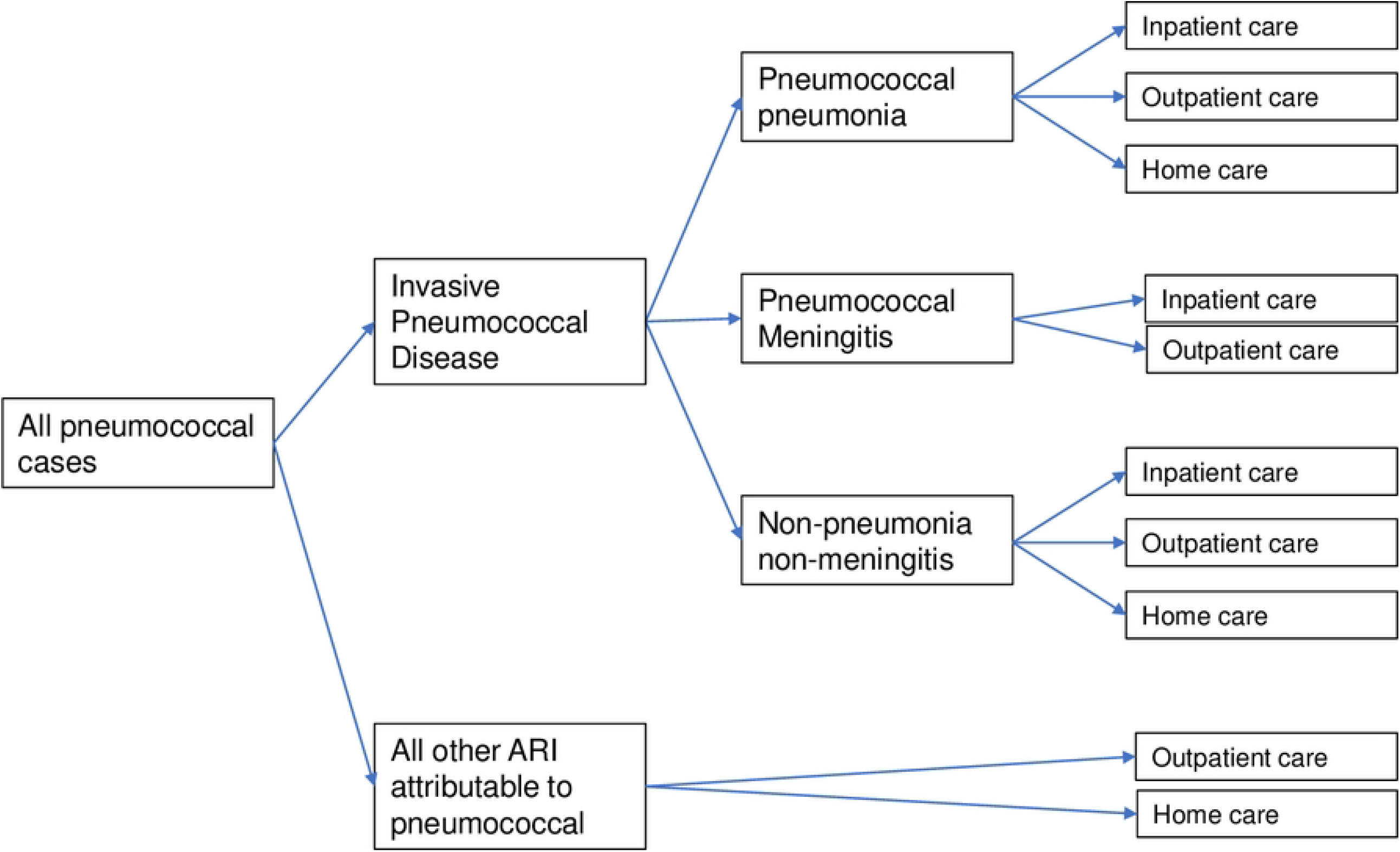
Health outcome tree of pneumococcal presentations and types of care.

Cases were modelled as receiving outpatient care, inpatient care, and at-home care (Table 2). Outpatient costs were estimated using WHO-CHOICE estimates for Sierra Leone [38]. The distribution of outpatient visits across facility type was informed by Sierra Leone’s 2019 Demographic Health Survey (DHS) [14]. We estimated inpatient costs using a Ghanaian costing study of pneumococcal pneumonia and meningitis, since WHO-CHOICE estimates of inpatient costs cover only hospital bed day costs. Ghana and Sierra Leone are both Western African nations which lie within the pneumococcal meningitis belt and have an established PCV schedule using three primary doses without a booster (3p+0). The Ghanaian costing study calculated a broader range of direct medical cost including the cost of medication, diagnostic tests, and hospital staff salaries in addition to hospital bed days. Costs for NPNM IPD were taken from pneumococcal pneumonia costs, as per Ojal et al. [21]. Home care costs were also taken from the Ghanaian costing study as WHO-CHOICE does not estimate these types of cost.

**Table 2.** Proportion of cases with access to health care and health system costs per event of care with point estimates, distributions, and sources. All costs presented in 2020 USD.

| Parameter | Component | Base estimate | Distribution | Reference |
| --- | --- | --- | --- | --- |
| Access to care | Outpatient care | 85.7% | Beta(84,14) | [14] |
|  | Home care | 14.3% | 1- Beta(84,14) |  |
|  | Inpatient care for IPD (general) | 65.2% | Beta(60,30) | [39] |
|  | Inpatient care for pneumococcal meningitis | 76.1% | Derived from inpatient access/outpatient access | [39] |
| Inpatient costs | Pneumococcal meningitis | \$172.57 | Uniform(137.71,396.74)* | [39] |
| | Pneumococcal pneumonia, and NPNM | \$157.16 | Uniform(140.16,405.79)* | [39] |
| Outpatient costs | All presentations | \$0.95 | Gamma(1.6,0.59) | [14, 38] |
| Home care costs | All presentations | \$1.59 | Uniform(0.00,3.55)* | [39] |

\* Distribution as per correspondence with Ghanaian study author [39]

Access to outpatient and inpatient care were modelled using care-seeking estimates from Sierra Leone’s DHS and hospitalisation rates from the Ghanaian study. Patients who did not have access to outpatient care were assumed to receive home care. We assumed that all cases of pneumococcal meningitis require hospitalisation (inpatient care), however not all had access to hospital care [1, 39]. We assumed that non-invasive pneumococcal disease (all other ARI attributable to pneumococcal) did not require hospitalisation.

### 2.6 Sensitivity analysis

We conducted both probabilistic and one-way sensitivity analyses to explore the characteristics under which a maternal pneumococcal vaccine would be cost-effective in reducing infant pneumococcal disease. Probabilistic sensitivity analysis was conducted by Monte Carlo simulation, that is, by running the model 1000 times randomly drawing per-patient from the probability distributions of all stochastic variables. We assumed that vaccine costs were fixed by government agreements and did not vary between individuals. Hence, we explored the effects of varied vaccine cost using one-way analysis.

## 3. Results

We estimated the incremental cost-effectiveness of a maternal vaccine program to be $16,145 per life saved, corresponding to $570 per DALY averted (Table 3). The cost of delivering the vaccine program ($1,022,305 per 100,000 infants) had a greater influence compared to health system costs averted ($69,376 per 100,000 infants) over the cost-effectiveness of the vaccine. Most vaccine program costs (82.2%) were attributable to the cost of the vaccine (Table 4).

**Table 3.** Cost per outcome averted under base model assumptions with 95% prediction interval from probabilistic sensitivity analysis. All costs presented in 2020 USD.

| Outcome | ICER per outcome | 95% prediction interval | Standard deviation |
| --- | --- | --- | --- |
| Life saved | 16,145 | 15,967-16,323 | 91 |
| DALY averted | 570 | 558-582 | 6 |
| Hospitalisation averted | 3,800 | 3,184-4,417 | 314 |
| Case averted | 1,006 | 994-1,017 | 6 |

**Table 4.** Breakdown of maternal vaccine costs by program component. All costs presented in 2020 USD.

| Component | Cost per 100,000 infants |
| --- | --- |
| Vaccine price | 840,821 (82.2%) |
| Vaccine dose wastage | 42,987 (4.2%) |
| Vaccine freight cost | 38,783 (3.8%) |
| Injection equipment (including wastage) | 4,716 (0.5%) |
| Operational costs | 94,998 (9.3%) |
| Total program cost | 1,022,305 (100.0%) |

Sensitivity analysis identified health outcome discounting rates, vaccine price and vaccine effectiveness to be highly influential over cost per DALY averted (Fig 2). Without discounting, the cost per DALY averted was $277 (53% of Sierra Leone’s GDP per capita). The cost per DALY averted would be $128 (24% GDP per capita) if PPV could be procured at the same cost relative to PCV in Sierra Leone as on the PAHO reference price list. The vaccine would be cost-effective under current market prices with an effectiveness of at least 81%.

**Fig 2.**
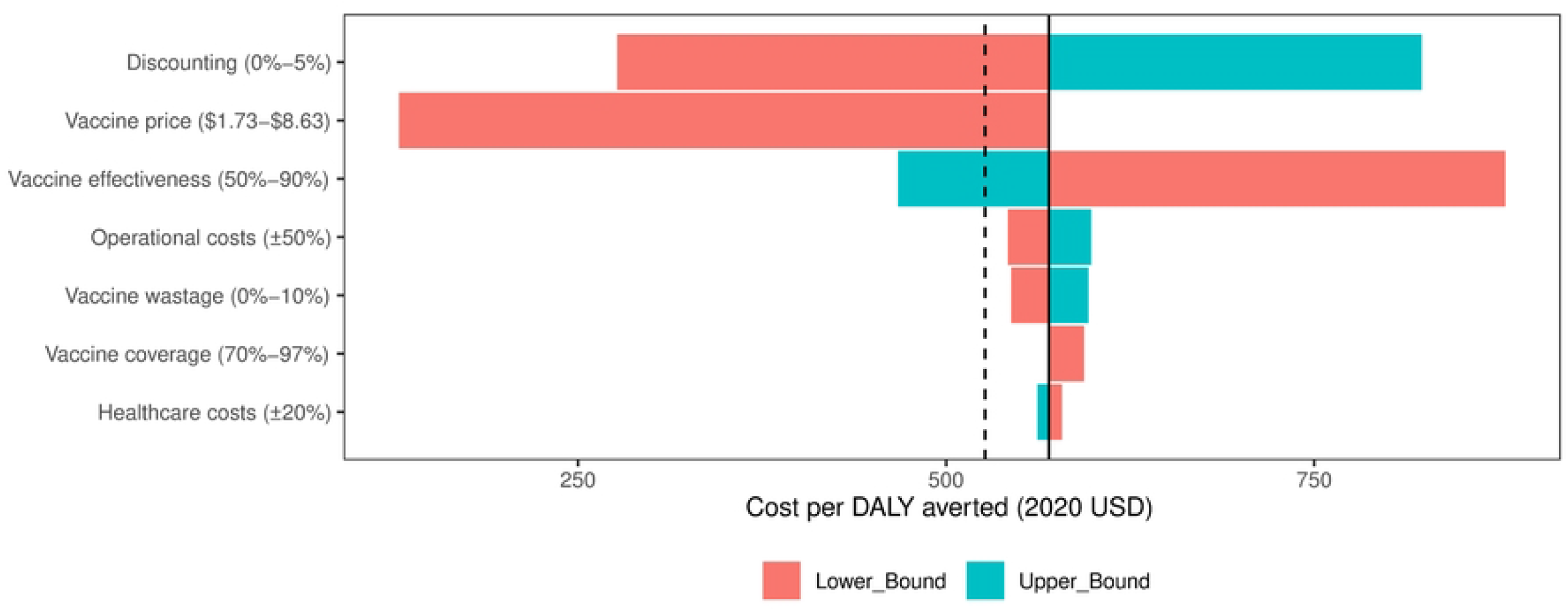
Tornado diagram representing one-way sensitivity analysis. The influence of key model parameters on incremental cost-effectiveness per DALY averted (2020 USD). Dashed line represents Sierra Leone’s GDP per capita [11].

## 4. Discussion

With current market prices, a maternal pneumococcal vaccine would not be cost-effective in Sierra Leone, employing Sierra Leone’s GDP per capita of $527 (2020 USD) as a benchmark [11]. We identify vaccine price as the largest cost driving the cost-effectiveness of a maternal PPV. This finding aligns with previous studies on the cost-effectiveness of new and underused vaccines, such as human papilloma virus (HPV), in low-income settings [40, 41].

The implementation of a cost-effective maternal pneumococcal vaccine would require negotiations with vaccine suppliers on vaccine price. The PAHO PPV reference price is two thirds of PAHO’s PCV reference price [28]. If a comparative PPV price were negotiated in Sierra Leone, a maternal pneumococcal vaccine would be cost-effective, $129 per DALY averted (23% GDP per capita) and $2.09/child. Further, an effective maternal PPV could significantly increase the global demand for PPV, as only ∼20 million doses are currently purchased annually [32]. Vaccine prices generally drop after introduction, especially through the increase in demand and involvement of humanitarian bodies such as GAVI [41].

Conventional methods of discounting the health effects of vaccines are being increasingly challenged [42-44]. In their critical review, Jit & Mibei establish that cost-effectiveness analyses of vaccines are particularly sensitive to discounting due to their distinctive characteristics compared to other health interventions [45]. Indeed, in this paper we demonstrate that the application of conventional 3% discounting shifts the cost-effectiveness of a maternal pneumococcal vaccine from $277 to $570 per DALY averted (from 53% to 108% of Sierra Leone’s GDP per capita). Hence, social values determining the choice of discounting rates may decide whether a maternal pneumococcal vaccine is cost-effective.

A limitation of our model was that we did not include all non-invasive pneumococcal presentations, such as otitis media and non-invasive pneumonia. Otitis media, in particular, causes a high burden of DALYs in Sub-Saharan Africa [46]. Additionally, we based hospitalisation data on utilisation from DHS surveys. Given the limitations in access to hospitalisation in Sierra Leone, this is likely to under-estimate actual disease-related health care utilisation if access was adequate. Consequently, our model underestimates the health system costs averted by a maternal vaccine.

Operational costs for maternal tetanus toxoid (TT) vaccination may be an overestimation of future operational costs for a maternal pneumococcal vaccine. The tetanus toxoid vaccine is the only maternal vaccine currently included in Sierra Leone’s Expanded Programme of Immunisation (EPI) [13]. We expect that the inclusion of an additional vaccine for an existing target group to be less costly. A recent systematic review identified that limited costing studies have been published detailing the costs of delivering maternal immunisation to pregnant women, particularly in low-income settings [35]. Future clinical trials should consider including a costing study component to better estimate the true operational costs of a maternal pneumococcal vaccine.

While our model provides a reasonable estimate of the cost-effectiveness of a routine maternal pneumococcal vaccine, our analysis does not include costs of introduction such as the expansion of cold chain storage capacity, and updates to vaccination cards. We expect respective Ministries of Health and Finance to undertake more detailed planning specific to their setting before introduction. Botwright et al.’s systematic review estimates HPV introduction costs to be 46% of total financial costs, and 32% of economic costs in the first year of program delivery [47]. Notably, previous vaccine introduction costs in Sierra Leone have relied upon GAVI funding [33, 48, 49].

Maternal pneumococcal vaccines demonstrate the potential to be cost-effective in low-income settings; however, their introduction will require negotiations with a vaccine provider or funding support from a humanitarian body. The current market price for PPV is too high for a maternal pneumococcal program to be cost-effective in our study setting of Sierra Leone. Further, the future use of a maternal PPV would require advanced planning with suppliers to avoid supply shortages.

## Data Availability

All relevant data are included within the manuscript.

## Acknowledgements

We acknowledge that this study would not have been possible without the efforts of previous studies used to inform our costing.

